# Validation of a transcriptome-based assay for classifying cancers of unknown primary origin

**DOI:** 10.1101/2022.05.06.22274683

**Authors:** Jackson Michuda, Alessandra Breschi, Joshuah Kapilivsky, Kabir Manghnani, Calvin McCarter, Adam J Hockenberry, Brittany Mineo, Catherine Igartua, Joel T Dudley, Martin C Stumpe, Nike Beaubier, Maryam Shirazi, Ryan Jones, Elizabeth Morency, Kim Blackwell, Justin Guinney, Kyle A Beauchamp, Timothy Taxter

**Affiliations:** Tempus Labs, Inc.

## Abstract

Cancers assume a variety of distinct histologies and may originate from a myriad of sites including solid organs, hematopoietic cells, and connective tissue. Clinical decision making based on consensus guidelines such as NCCN is often predicated on a specific histologic and anatomic diagnosis, supported by clinical features and pathologist interpretation of morphology and immunohistochemical (IHC) staining patterns. However, in patients with nonspecific morphologic and IHC findings—in addition to ambiguous clinical presentations such as recurrence versus new primary—a definitive diagnosis may not be possible, resulting in the patient being categorized as having a cancer of unknown primary (CUP). Therapeutic options and clinical outcomes are poor for CUP patients, with a median survival of 8-11 months. Here we describe and validate the Tempus Tumor Origin (Tempus TO) assay, an RNA-seq-based machine learning classifier capable of discriminating between 68 clinically relevant cancer subtypes. We show that the Tempus TO model is 91% accurate when assessed on retrospectively and prospectively held out cohorts of containing 9,210 samples with known diagnoses. When evaluated on a cohort of CUPs, the model recapitulated established associations between genomic alterations and cancer subtype. Combining diagnostic prediction tests (e.g., Tempus TO) with sequencing-based variant reporting (e.g., Tempus xT) may expand therapeutic options for patients with cancers of unknown primary or uncertain histology.

## Introduction

Accurate characterization of primary tissue origin, histology, and clinical/pathologic stage is required for assigning effective therapeutic interventions for cancer patients ^1^. However, some patients present with ambiguous clinical and histologic findings and no definitive primary site of disease ^2^. These tumors are a heterogenous group known as cancers of unknown primary (CUP) and comprise 2-5% of cancers ^3,4^. The American Cancer Society estimates that over 30,000 patients will be diagnosed with CUP in 2021 ^5,6^. For CUP patients, the average survival time is typically 8-12 months after diagnosis, though some subgroups may survive for up to 12-36 months ^4^. Establishing why certain CUP patients have more favorable prognoses and improving survival for all CUP patients is a key goal for clinicians ^7,8^.

Although direct examination of tissue—via morphology and immunohistochemistry—has long guided cancer type diagnosis, advances in sequencing technologies have facilitated new ways of characterizing cancer ^9,10^. These approaches have further enabled the development of several molecular diagnostics aimed at determining tumor origin specifically ^11^. For example, one commercially available assay leverages RT-PCR of 92 genes and a machine learning algorithm to differentiate between 50 tumor subtypes ^12–15^. Additional methods for tumor classification rely on microRNA signatures ^16,17^, transcript expression ^18–20^, mutation profiling via DNA sequencing ^21,22^, methylation profiling ^23^, and whole-slide histology imaging ^24^.

While prior studies and assays have made important advances, they also have a number of limitations ^11^. For instance, DNA-based panels ^21^ perform well when a tumor has a canonical alteration associated with a specific cancer type or subtype, but fail to accurately identify the subtype when no such mutations exist. RT-PCR assays that profile a subset of the transcriptome may have limited accuracy when a tumor does not express a lineage-specific gene ^14,15^. Many assays can predict tissue of origin, but cannot predict site-specific subtype or histology; this lack of specificity may be inadequate for some treatment or diagnostic decisions. Multi-omic machine learning architectures that integrate data from multiple assays may provide an opportunity for accuracy improvement and can help alleviate concerns with using single data types ^25^, but these models require complex algorithms for data integration. Finally, even IHC—the current standard of care in making CUP diagnoses—is limited by the frequent loss of cell markers during de-differentiation and oncogenesis ^26^.

By contrast, full-transcriptome RNA-seq overcomes several of these limitations. As shown in prior work using methylation ^23^ and gene expression ^14,15^ models, cancer cells retain an epigenetic and transcriptional signature of their cell type of origin. Next-generation sequencing (NGS) has facilitated the adoption of transcriptome-wide expression profiling in a clinical setting owing to relatively low costs and minimal sample preparation. Hybrid capture assays, with probes covering the whole exome, can accurately extract expression profiles from small formalin-fixed paraffin-embedded (FFPE) tissue biopsies. Expression data generated by transcriptome-wide RNA-seq are well-suited for machine learning and statistical modeling due to their quantitative, high dimensional, and untargeted nature ^27^.

Here, we describe and validate an RNA-expression-based tumor diagnosis classifier trained on whole-exome capture RNA-seq data from 43,726 tumor samples. This machine learning model distinguishes 68 tumor subtypes—a more comprehensive and finer level of subtype resolution than previous studies ^14,21,25^. These tumor subtypes include neuroendocrine subtypes, sarcoma subtypes, and site-specific histologies to allow for precise application of medical guidelines. The classifier is highly accurate (91%) on an independent validation dataset and is robust to primary versus metastatic lesions and tumor purity.

## Materials and methods

### IRB

All analyses were performed using de-identified data; IRB exemption Pro00042950 was obtained from Advarra on April 15, 2020.

### Cohort

Cohort selection for training and validation is summarized in **Fig. 1**. The principal inclusion criterion was the availability of quality-controlled RNA sequencing of a tumor specimen. A summary of assay quality controls is described in the “RNA-seq assay” section. No restrictions on the cancer stage or site of biopsy were applied. Biospecimen material was restricted to FFPE, bone marrow, and blood. Allowed cancer types and subtypes are described in section “Cancer subtype assignment”. Samples were either labeled with known subtype (cancers of known primary) (N=52,936, comprising 68 subtypes) or marked as unlabeled (e.g., CUP) (N=1,708). Labeled samples were split into 75%/25% training and validation cohorts via stratified random sampling (matching subtype distributions), which were used for model training and evaluation, respectively. The validation cohort was an independent test set of tumors derived from different patients and samples than the training data. A comparison of the tissue and subtype distributions in the training and validation sets is provided in **Supplementary Tables S1** and **S2**. In the event that a patient had multiple biopsies, they were all grouped into either the training or validation cohorts to prevent information leakage. The validation cohort consisted of both retrospective samples that were sequenced prior to the model training date, as well as prospective samples sequenced after model training. The unlabeled (CUP) cohort consisting of diagnostically ambiguous tumors was not used in the primary performance analysis, but was analyzed as part of the mutation-subtype enrichment study.

**Figure 1.**
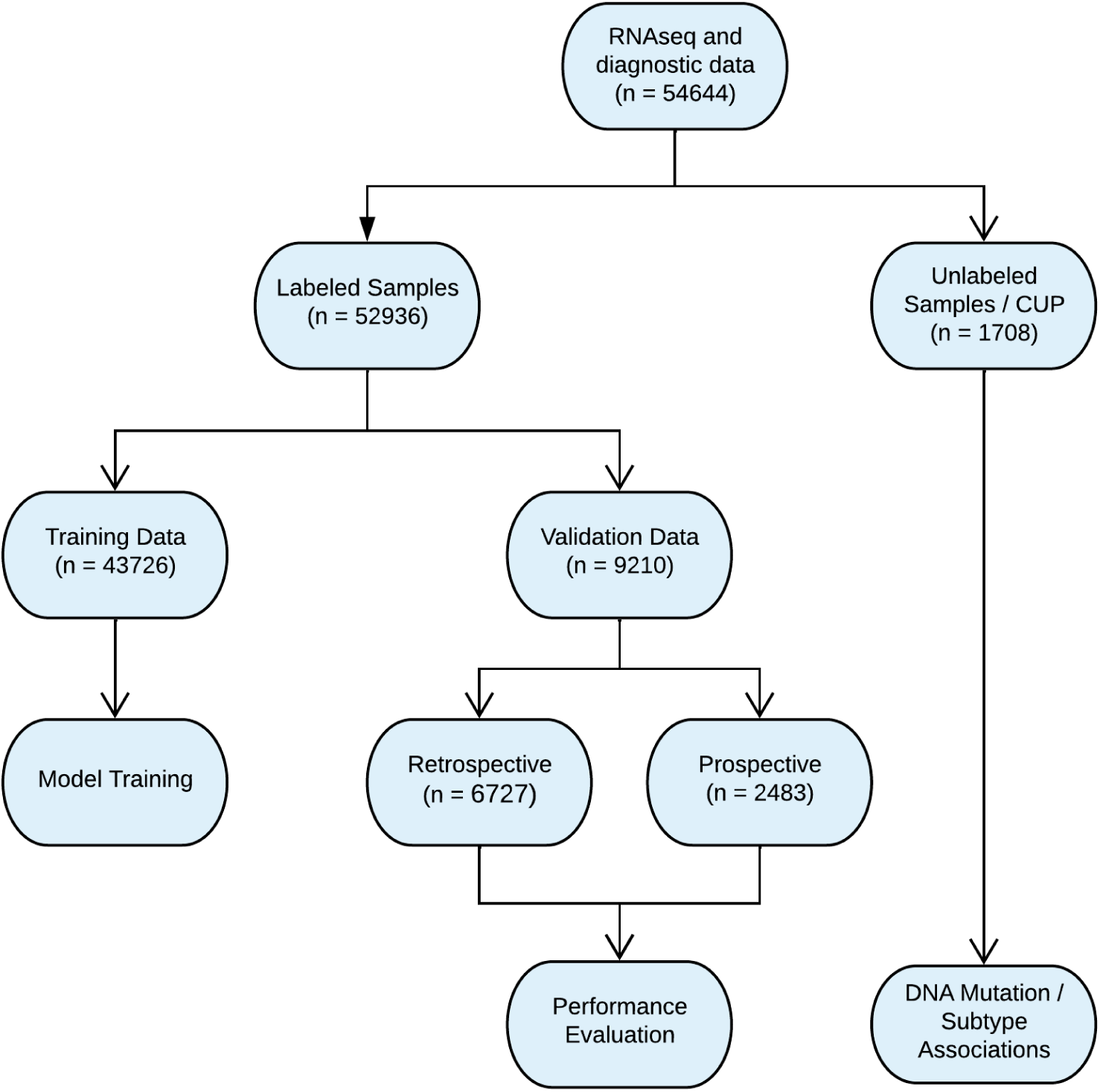
CONSORT diagram describing the cohorts used for classifier training, accuracy evaluation on labeled samples (validation data), and assessment on unlabelled CUP samples.

### RNA-seq assay

As part of the Tempus xT next-generation sequencing assay, CAP/CLIA validated hybrid-capture RNA-seq was used to generate transcriptome-wide expression data from paraffinized tissue ^28,29^. Briefly, FFPE samples were cut and reviewed by a pathologist and samples of adequate quality—with a minimum tumor fraction of 20% after macrodissection—underwent RNA extraction. Extraction quality was assessed using a fragment analyzer, and quantity was assessed using a fluorescent nucleic acid stain and a fluorescence microplate reader. At least 50 ng of RNA were extracted before proceeding to library preparation. Library preparation included steps for complementary strand synthesis with reverse transcriptase, ligation of dual indexed unique molecular identifier (UMI) adapters, and PCR amplification. At least 150 ng of amplified cDNA were required to proceed to hybridization. cDNA fragments were captured using a full exome panel ^30^ and sequenced on a NovaSeq 6000 (Illumina Inc, San Diego, CA) to a minimum depth of 30 million reads.

Following sequencing, raw BCL files were used to generate FASTQ files using BCL2FASTQ (v2.17). Adapters were trimmed using Skewer (v0.2.2). Reads were then aligned with STAR (v.2.5.4a) to generate BAM files and undergo UMI deduplication with umitools (v1.0.1). Following deduplication, samples were converted to FASTQ files using bedtools (v2.27.1) before undergoing quantification using Kallisto (v0.44.0) with the ensembl GRCH37 reference transcriptome ^28,29^.

### Feature engineering

Raw RNA expression data were normalized to minimize the effect of technical artifacts such as GC-content, transcript length, and library size ^28,29^. This normalization strategy is similar to the one employed in DESeq ^31^, but adjusted to generalize the transformation to apply to new samples not included in the training set (see Supporting Information). In addition, a batch correction method was used to account for small technical differences between two versions of the RNAseq assay, including a modified exome-capture probe set and the addition of UMIs. The batch correction method leverages more than 500 paired samples that were run on both assays to match the means and variances of normalized expression (see Supporting Information for details). In this work, all validation statistics refer to the most recent assay version.

The gene-level expression features underwent several threshold-based filters and data scalings. First, low variance genes were removed. Second, genes with low inter-assay correlation were removed. Finally, expression data were scaled to have a uniform mean and variance per gene. The optimal thresholds for establishing low variance and inter-assay reproducibility filtering were tuned via a hyperparameter grid search, as described in detail below. To prevent information leakage, all transformation parameters were learned using the training set and independently applied to the validation set.

### Expression validation

Gene expression features were analytically validated ^32^ to ensure that the model relied on accurate and reproducible inputs. Universal Human Reference (UHR) RNA ^33^ was sequenced on the Tempus RNA-seq assay to establish linearity against an orthogonal method. Normalized gene expression values were compared against a reference set of 17,321 qPCR ΔCT values established by the MAQC consortium. Normalized expression data were generated for 21 UHR replicates. All replicates had a Pearson’s correlation coefficient (*R* value) greater than 0.76 between the ΔCT values and gene expression data. Further methodological validation included a per-gene linearity study using 88 clinical samples and testing 18 genes to measure concordance between qPCR ΔCT values and normalized gene expression levels (**Supplementary Fig. S1**). Of these, 15/18 genes had an *R* value > 0.75. All genes had an *R* value greater than 0.5, and the lowest performing genes were those with a small ΔCT dynamic range. In genes with large dynamic range, gene expression profiling with RNA-seq is highly concordant with qPCR.

### Diagnostic subtype assignment

Samples were annotated with one of 68 subtypes (in this work, a subtype is defined as a site-specific histology) inclusive of cancer types and histologic subtypes. Samples were annotated with clinical and diagnostic information in two ways. Each sample underwent pathology review by a board-certified pathologist. The pathologist workflow included reviewing a patient’s clinical documents (such as progress notes and external pathology reports which includes diagnostic IHC), viewing H&E stained images, and inputting free-text diagnostic data describing the primary site, histological subtype and biopsy site into the lab information management system for a sample. Patient records underwent a second round of clinical data review by trained abstractors to generate standardized diagnostic data for a patient. Abstractors reviewed a patient’s clinical notes within a document review platform and assigned Unified Medical Language System codes to describe a patient’s clinical history of diagnosis.

From an analysis of this corpus of real-world diagnostic data, a curated set of diagnoses was developed to achieve broad coverage over, and clinical differentiation of, clinically meaningful cancer subtypes and histologies. In total, 68 diagnostic labels (**Supplementary Table S3**) were developed. To assign diagnostic labels to each sample, a natural language model consisting of regular expressions was used to parse the free text diagnostic field assigned by the pathologist into a label. A rules-based system was designed to identify diagnostically ambiguous cases where a definitive subtype could not be confidently assigned. Diagnostically ambiguous samples (e.g., samples matching zero or several subtypes) were excluded from the labeled model training and validation sets, but were considered in the analysis of the CUP cohort.

De-identified case review by pathologists was used to assess the accuracy of automated subtype assignments. Three different pathologists were presented with de-identified clinical data from 118 cases (1 case from each of the subtypes and 50 additional cases selected at random). Each pathologist reviewed the clinical data and was instructed to assign a single subtype to each case from the set of 68 labels.

### Machine learning model

The machine learning model is a multinomial logistic regression classifier with L2 regularization. To overcome class imbalance in the training set, rare classes were upweighted. The weights for the *i-*th class are given as 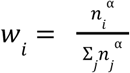, where *n* is the observed count of class *i* and *α* is a smoothing parameter that was determined via hyperparameter search. Including the previously mentioned feature engineering, the hyperparameter grid search was performed via 3-fold cross-validation (evaluated using the log loss metric) on the training set that simultaneously assessed: 1) L2 regularization strength, 2) variance threshold, 3) interassay concordance threshold, and 4) class weight smoothing parameter.

### Validation metrics

Models were evaluated using three metrics computed on the independent validation set: accuracy, top-3 accuracy, and mean sensitivity. Accuracy and top-3 accuracy were computed as the average (over samples) of the number of times the true subtype assignment was found in the top one or any of the top three highest probability subtype predictions, respectively. Mean sensitivity was computed by first calculating the sensitivity [TP/(TP+FN)] within each subtype, and then taking the unweighted mean across all 68 subtypes. Binomial confidence intervals for metrics were estimated using Markov chain Monte Carlo. Per-label specificity [TN/(TN+FP)] was also computed within each subtype.

### TCGA assessment

To assess the generalizability of our model beyond samples sequenced in our laboratory, we evaluated the performance of the TO classifier using data generated by The Cancer Genome Atlas (TCGA) ^34^.

First, tabular clinical data for all TCGA studies were downloaded from the GDC data portal (https://portal.gdc.cancer.gov/). Next, a crosswalk was constructed to map each unique TCGA subtype (*i*.*e*., each of the 617 unique combinations of TCGA study, ICD10 code, primary diagnosis, site of resection, and tissue of origin) to the appropriate Tempus TO subtype (**Supplementary Table S4**). Subtypes with conflicting diagnostic information or no Tempus equivalent were labeled “Other” and excluded from analysis. Overall, 33 TCGA studies were mapped to 38 Tempus TO subtypes (**Supplementary Table S5**). One notable example of the histology-by-histology mapping was TCGA type “sarc”, which was mapped onto 5 distinct Tempus TO sarcoma subtypes. Another example is the TCGA typing for colon (“coad”) and rectal (“read”) carcinomas, which were mapped to Tempus TO subtype “colorectal adenocarcinoma”; this lumping is consistent with analysis of these TCGA types presented in the TCGA publication ^35^, which combined the types during analysis.

After obtaining the crosswalk, the diagnostic subtypes specified by TCGA were mapped to obtain a comparable set of internally-defined subtype labels for each TCGA sample. The RNA sequencing results from TCGA fastqs (9,976 samples) were processed using Kallisto and normalized at the gene level with the same reference for consistency with other data used in this work. Finally, the subtype classifier was applied to each sample in TCGA to obtain a predicted subtype and performance was assessed using the same metrics described above.

### DNA sequencing assay

Each sample also underwent co-isolation of nucleic acids to generate both DNA and RNA material for sequencing. DNA was sequenced using a targeted sequencing panel (Tempus xT) ^29^. The assay has an average coverage of 500x and detects single nucleotide variants, indels, and copy number variants in 595-648 genes spanning 3.6 Mb of genomic space. DNA data was not used as input into the gene expression-based model, so it provides an orthogonal method to aid in diagnostic interpretation and for evaluation of self-consistency on CUP samples.

### Mutation-subtype associations

Mutation-subtype enrichments were used to characterize classifier behavior on CUP samples without a definitive clinical diagnosis. First, somatic mutations were aggregated for two cohorts: samples with a subtype diagnosis and samples without (CUP samples). For samples with a known subtype, Fisher’s exact test was used to find all subtype-gene pairs with significant enrichments. A significant enrichment was defined as an association (*i*.*e*., an odds ratio calculated from the contingency table between subtype and mutation status) with a one-sided p-value less than 1.6e-6 (i.e., an alpha of 0.05 using the Bonferroni-correction with 30,804 comparisons, obtained by considering 453 genes with observed mutations for one of the 68 labels). Significant enrichments were further filtered based on their frequency; subtype-gene pairs with insufficient statistical power in the CUP cohort were removed by excluding pairs for which the probability of observing zero mutant counts (assuming identical mutant frequencies across labeled and CUP cohorts) was below 0.05. Finally, the list of significant associations (from known subtypes) was then evaluated on the classifier-predicted subtypes in the CUP cohort. Somatic mutation frequencies were estimated as the fraction of samples with a mutation in each gene.

## Results

### Label Accuracy

We compiled an initial dataset containing 54,644 tumor-derived RNA-seq samples from the Tempus database (**Fig. 1**). Approximately 3% of these samples (N=1,708) were from cancers with an unknown primary site of origin (CUPs), which were withheld from model development and used only for downstream validation and model interrogation. For the remaining samples (N=52,936), we assigned one of 68 subtype labels based on clinical documentation and a standardized abstraction protocol (see Diagnostic subtype assignment in Materials and Methods).

To evaluate the appropriateness of the cancer subtype labels for tumor origin classification, three pathologists assessed 118 randomly-selected, blinded, de-identified cases. The pathologists were then asked to assign a label to each case. For 109/118 cases (92%), the three pathologists agreed in their assignment. Treating those “consensus pathologist” annotations as the gold standard, the diagnostic subtype assignment method was 98% accurate (107/109). On the full 118 sample dataset, the concordance of each pathologist with the automated assignment was 92% (109/118), 94% (111/118), and 97% (114/118).

### Model performance on labeled samples

We applied the trained Tempus TO model to the independent validation dataset to assess overall performance. As the Tempus TO model outputs a probability that a sample belongs to a given label, we first calculated performance based on the highest probability label assigned to a sample (**Table 1**). The model accuracy was 91.1% (95% CI: 90.5% - 91.6%); this number approximates the probability that a random sample (with known subtype) from our laboratory is correctly predicted by the RNA-based classifier. As *accuracy* overemphasizes the most common cancers in our dataset, such as colorectal adenocarcinoma, we also assessed performance using *mean sensitivity*, which balances the impact of rare and common subtypes (see Material/Methods). In the validation cohort, we achieved a mean sensitivity of 80.0% (95% CI: 77.9% - 81.7%). Sensitivity per subtype ranged from 25.9% (small bowel adenocarcinoma) to 100% in ependymoma, ewing sarcoma, meningioma, renal chromophobe carcinoma, and schwannoma (**Fig. 2**). Full results of all metrics for each subtype are listed in **Supplementary Table S3**.

**Table 1.** Model performance metrics on a held-out set of labeled samples.

|  | validation |
| --- | --- |
| Sample Size | 9210 |
| Accuracy | 91.1% |
| Top 3 Accuracy | 97.5% |
| Mean Sensitivity | 80.0% |

**Figure 2.**
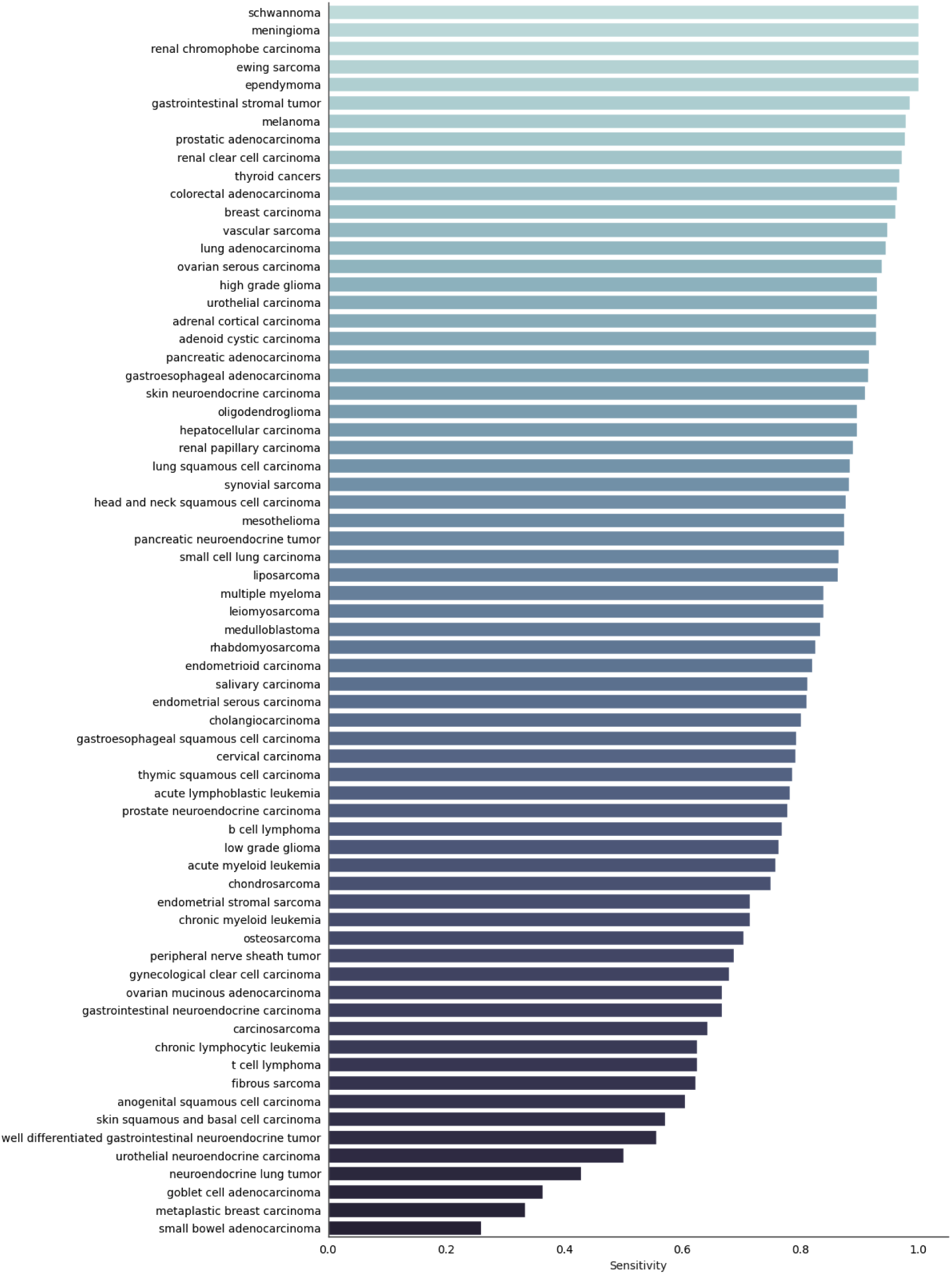
Sensitivity for each of the 68 possible subtypes sorted from highest to lowest. See **Supplementary Table S3** for full results.

Since our classifier outputs a probability that a sample belongs to one of 68 different subtypes, we hypothesized that mislabeled samples might be correctly identified within the top N predicted subtypes. When considering the top N predicted subtypes—*i*.*e*., a prediction is deemed correct as long as the correct label is amongst the top N predictions^36^—we observed considerable increases in overall accuracy. Crucially, for subtypes that were most difficult to predict, the correct prediction was frequently the second highest predicted subtype, and many subtypes are predicted perfectly when considering the top 3 highest predictions (**Fig. 3**). Overall, this shows that when the model makes an incorrect prediction, the correct prediction is very often either the next highest prediction or among the top 3.

**Figure 3.**
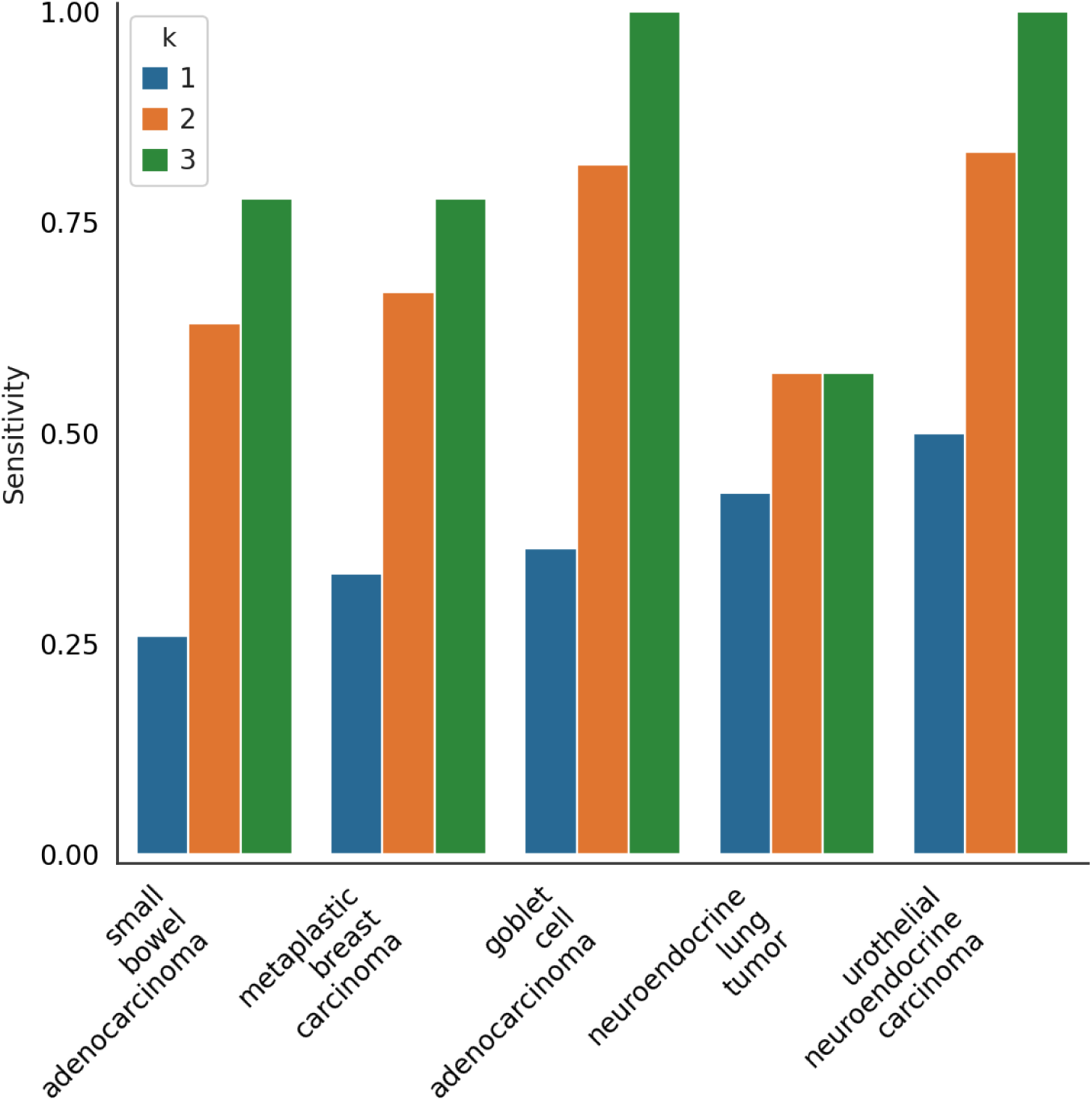
The top-k sensitivity (k=1,2,3) for the most difficult to predict subtypes highlights that the correct prediction is frequently in the top-2 or top-3 predictions.

To assess the possibility of model drift and to ensure model generalizability, we examined accuracy separately across the retrospective (data that was available at the time of training but computationally withheld) and prospective (data that was sequenced after model development) sub-cohorts (**Supplementary Table S6**). We found no significant difference in accuracy (91.2% and 90.8%, respectively; Fisher’s exact test, *p*=0.6), further indicating that the model is generalizable. Model performance was found to be robust to tumor purities down to 10%, showing an accuracy of at least 89% for all purity bins above 10% (**Supplementary Table S7**). Further, there was only a small reduction in performance when applied to cases with imputed metastatic, non-metastatic, and unknown primary status—as defined via curated sample metadata and pathological review (**Supplementary Table S8)**.

While overall metrics of model accuracy are critical for assessing performance, these single numbers obscure the actual patterns of correct and incorrect predictions. We therefore examined confusion matrices (predicted vs observed subtype counts) for several clinically relevant groupings of subtypes (**Supplementary Fig. S2**). As expected, there are molecular similarities between less common gastrointestinal subtypes like goblet cell adenocarcinoma and small bowel adenocarcinoma and more common subtypes such as colorectal adenocarcinoma. Furthermore, the confusion matrices demonstrate robust per-label sensitivity across liver and lymph node tissue sites.

Lastly, we note that a small percent of predictions yield possibly ambiguous results. These samples are flagged (and no-called) whenever the largest predicted probability (the max(p) value) from among the 68 possible labels is below 35%. Among validation set samples, this occurs approximately 1% of the time. Samples passing the max(p) threshold have a marginally higher accuracy (91.7%). With the exception of this paragraph, we report all metrics according to the more pessimistic scenario where the largest predicted probability is reported regardless of whether it is below 35%. In clinical applications of the model, however, samples with max(p) below 35% will be reported as indeterminate.

### Generalization to TCGA Samples

To assess the generalizability of the classifier beyond samples sequenced in our laboratory, we performed an analysis of subtype-labeled samples obtained from TCGA. Despite the classifier never having seen TCGA data and the possibility of technical batch effects based on differences in RNA sequencing protocols, the overall accuracy of the TO classifier was 84.3% on TCGA samples and the mean sensitivity was 85.2%—both values are on par with the reported accuracy from our retrospective and prospective validation sets. Full TCGA performance metrics are shown in **Supplementary Tables S9** and **S10**, with full confusion matrices for Tempus and TCGA shown in **Supplementary Figs. S3** and **S4**.

### Mutation-subtype associations in CUP sample predictions

The previous analyses were all performed on samples of known primary (since this is necessary in order to label samples for either training or testing), but true CUP samples may present particularly unique challenges. Assessing performance of the TO classifier on CUP samples–which, by definition, have no known subtype–is difficult but many DNA alterations are associated with specific cancer subtypes ^37^. While we note that DNA variant information is not diagnostically sufficient for establishing tumor subtype, we hypothesized that mutation-subtype associations in the DNA data of labeled samples should be similar to associations found among the predicted subtypes of CUP samples (see Methods). Because the Tempus TO model is fully blind to DNA sequence variant data during training and inference, this analysis provides an independent self-consistency characterization of the classifier in the CUP setting.

For each subtype and gene alteration (SNV, indels), we selected significant mutation-subtype associations, identifying 158 enrichments in the labeled cohort (see Methods); these significant associations involved 29 subtypes and 71 genes. We next asked whether these same associations were observed in CUP samples, and were able to recover positive subtype-gene associations (*i*.*e*., subtype-gene odds ratios > 1) in 150 of the 158 cases (94.9% of associations recovered; 95% CI: 91.5%–98.3%) despite our model having no explicit knowledge of sequence variants. Finally, the somatic mutation frequencies from the known subtype are recapitulated in CUP samples with the corresponding predicted subtype (**Table 2, Figure 4)**. Overall, this assessment found that TO predictions on CUP samples are able to recapitulate known mutation-subtype associations and is a further indicator of the consistency of the TO classifier.

**Table 2.** Observed somatic mutation frequency (in the labeled and CUP cohorts) of the ten most significant enrichments in the CUP cohort. See **Supplementary Table S11** for the full list.

| Subtype | Gene | Somatic Mutation Frequency |  |  |  |
| --- | --- | --- | --- | --- | --- |
|  |  | Labeled Cohort | CUP Cohort | Labeled Cohort | CUP cohort |
|  |  | Within Subtype |  | Outside Subtype |  |
| pancreatic adenocarcinoma | *KRAS* | 0.530 | 0.681 | 0.101 | 0.132 |
| lung adenocarcinoma | *STK11* | 0.092 | 0.241 | 0.007 | 0.019 |
| cholangiocarcinoma | *IDH1* | 0.079 | 0.109 | 0.011 | 0.002 |
| lung adenocarcinoma | *KEAP1* | 0.050 | 0.159 | 0.003 | 0.014 |
| colorectal adenocarcinoma | *APC* | 0.496 | 0.309 | 0.025 | 0.036 |
| lung adenocarcinoma | *SMARCA4* | 0.032 | 0.144 | 0.007 | 0.026 |
| small cell lung carcinoma | *RB1* | 0.436 | 0.365 | 0.035 | 0.053 |
| cholangiocarcinoma | *BAP1* | 0.075 | 0.117 | 0.007 | 0.017 |
| skin squamous and basal cell carcinoma | *NOTCH1* | 0.229 | 0.333 | 0.011 | 0.014 |
| skin squamous and basal cell carcinoma | *NOTCH2* | 0.111 | 0.273 | 0.006 | 0.005 |

**Figure 4.**
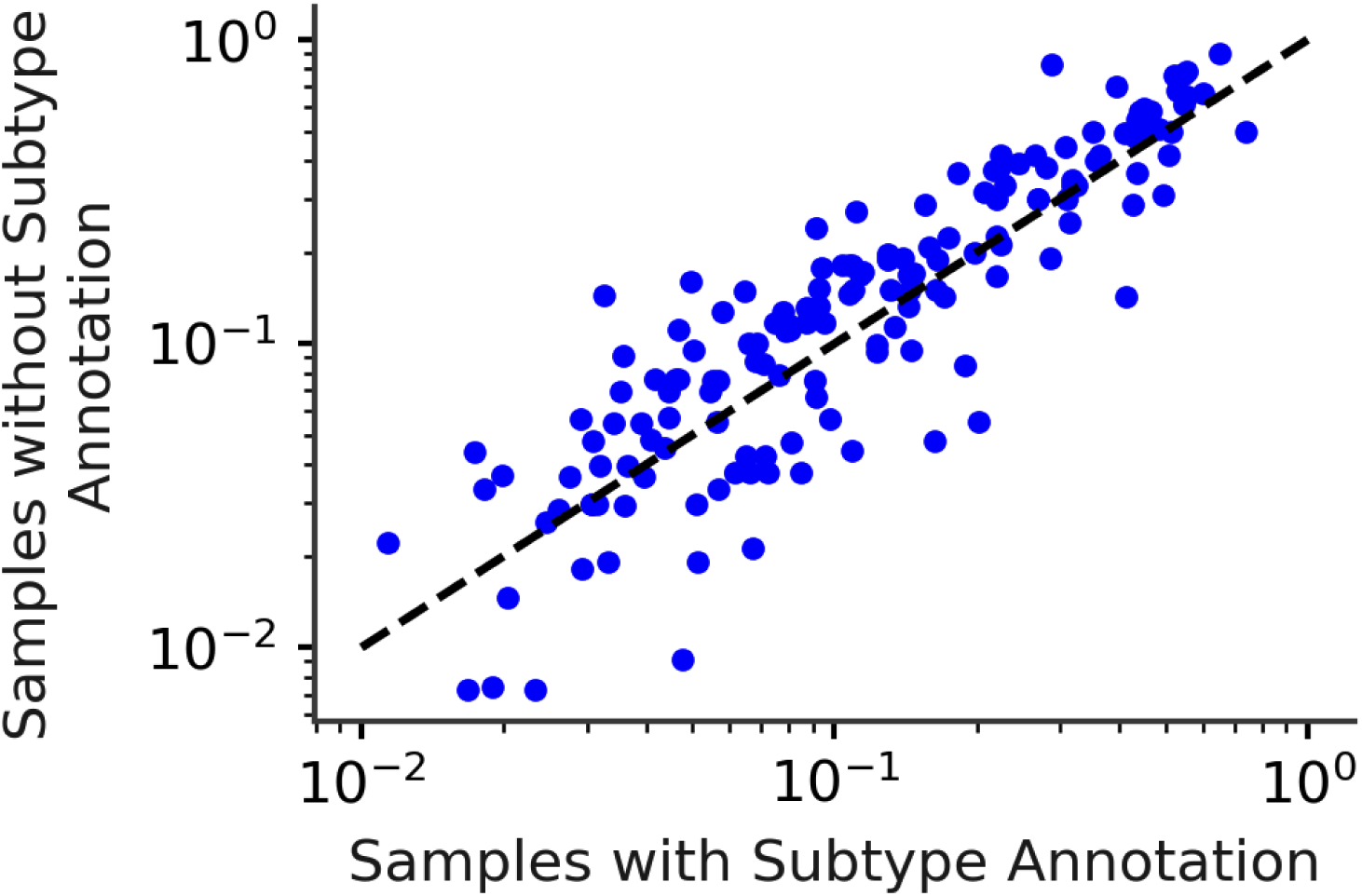
Somatic mutation frequencies are compared for tumors of known origin (labels obtained from clinical data) and tumors of unknown origin (labels predicted by the Tempus TO classifier). Each point represents the somatic mutation frequency for a particular gene and subtype; all gene-subtype pairs passing the labeled-cohort significance threshold are included.

## Discussion

Improving outcomes for CUP patients remains an unmet clinical need. Given the increasingly low-cost and widespread availability of genomic testing, machine learning approaches—such as the one developed and validated here—promise to improve clinical management for CUP patients by providing a specific anatomic and histologic diagnosis. We show that the Tempus TO assay achieves a 91% classification accuracy across 68 well-defined and clinically relevant tumor subtypes using only RNA expression data, which is quantified as part of the Tempus xT sequencing assay. To further validate the Tempus TO assay and ensure its robustness, we assessed a number of orthogonal metrics for measuring classification accuracy, ensured accuracy on independent retrospective and prospective cohorts, and finally showed that subtype predictions are capable of recapitulating subtype-specific mutational patterns. We additionally evaluated the TO model in TCGA data to demonstrate generalizability and found comparable performance to the retrospective and prospective validation sets.

An extension of the value that comes with the diagnostic resolution of CUPs provided by Tempus TO is the potential impact on therapeutic decision making as it relates to supporting NCCN recommended guidelines and FDA label indications in both non-biomarker and biomarker dependent contexts. The non-biomarker context highlights an application of Tempus TO beyond the traditional CUP setting to aid in the evaluation of tumors with a clear primary site of origin but conflicting or ambiguous histology. In lung cancer for instance, the choice to give bevacizumab in combination with carboplatin and paclitaxel is dependent on a differential diagnosis between lung adenocarcinoma and lung squamous carcinoma ^38^. However, lung tumors can present with poorly differentiated histology limiting further classification beyond “non-small cell lung cancer” ^39^ and complicating the ability to follow histology-specific guideline recommendations. Therefore, a critical component of any subtype classifier is the ability to discriminate between histologies. The Tempus TO assay has sensitivity to differentiate between lung squamous cell carcinoma (sensitivity: 0.88), lung adenocarcinoma (0.95), small cell lung carcinoma (0.87), and neuroendocrine lung tumors (0.43) with less tissue than is typically required for IHC (**Supplementary Fig. S2**). Judicious use of IHC in small tissue samples to determine a histological diagnosis is recommended to conserve tumor tissue, especially in patients with advanced disease or limited biopsy material ^40,41^.

Biomarker dependent contexts highlight the value of Tempus TO paired with targeted molecular profiling assays, such as Tempus xT, where diagnostic predictions can support biomarker and cancer type specific therapy indications. An example of this would be differentiating diagnostically challenging upper gastrointestinal neoplasms such as gastroesophageal carcinoma, cholangiocarcinoma, hepatocellular carcinoma, pancreatic adenocarcinoma, pancreatic neuroendocrine tumors, small bowel adenocarcinoma, gastrointestinal neuroendocrine carcinomas, and well-differentiated gastrointestinal neuroendocrine tumors (**Supplementary Fig. S2**). With the clinical approval of *FGFR2* ^42^ and *IDH1* ^43^ targeting therapies in cholangiocarcinoma, accurate diagnosis and mutational analysis of this rare tumor subtype has the potential to support on-label therapeutic options.

Despite the high accuracy of the Tempus TO model, there are nevertheless several caveats that we wish to emphasize as possible limitations and areas for ongoing research.

First, any diagnostic algorithm for CUP patients presents innate evaluation challenges since CUPs do not—by definition—have ground-truth labels to use in evaluation. Analytical performance can only be evaluated on cancers with an adjudicated diagnosis—calling into question the generalizability of model performance to the intended population of the assay. However, we partially addressed this challenge using DNA associations in the CUP population as an orthogonal assessment. A supporting example from the DNA analysis is the enrichment of *SMARCA4* alterations in lung adenocarcinomas (**Table 2**). *SMARCA4* is one of the catalytic subunits of the SWI/SNF chromosomal remodeling complex, a critical transcriptional regulator. *SMARCA4* lung cancers have been shown to be associated with poor histologic differentiation and a lack of TTF1 staining which is a commonly used IHC biomarker for diagnosing lung cancer^44^. Although *SMARCA4* mutations are not entirely specific for lung, our analysis finds that *SMARCA4* mutations are significantly associated with lung adenocarcinoma in both CUP and non-CUP settings—but with an almost five-fold higher somatic mutation rate in the CUP cohort. Therefore, our finding of enrichment of *SMARCA4* variants in CUP patients predicted as lung adenocarcinoma is an expected finding given the diagnostic challenge posed by TTF1 negative, poorly differentiated carcinomas ^45^.

Second, performance metrics such as overall accuracy are dependent on the case distribution observed in our laboratory, which may differ from other institutions as well as the general population. To mitigate the effect of differing case distributions, we additionally report per-label sensitivity and specificity for use in evaluating each label in isolation (**Supplementary Table S3**). It should be noted that one of the expected drivers of misclassification in the TO test is the overlapping nature of gene expression profiles for cancers with similar histologies such as squamous cell carcinomas from several primary sites, or cancers of differing histology but from the same site. Evaluation of confusion matrices that illustrate per-label performance identify similar trends of misclassification attributable to this expected limitation in both the Tempus and TCGA data (**Supplementary Figs. S3** and **S4**). Additionally regarding possible TCGA to Tempus per-label discordance, there were expected limitations in the mapping of TCGA and Tempus labels and different diagnostic practice variability such as with the evolving WHO classification of gliomas^46^.

Third, while our label set is comprehensive, it does not represent all possible cancer subtypes. In clinical practice, a CUP case may be presented to the classifier where the true diagnosis is not well represented in the 68 diagnostic cancer subtypes or is unlike any case observed in the training set. Examples of such cases may include rare *de novo* histologic variants of common cancers such as sarcomatoid variants. The resulting predictions in these settings can still inform the possible site of origin and the histologic subtype, however, oversight by the ordering physician is required to integrate molecular predictions with all available clinical evidence to aid in subtype diagnosis.

Fourth, this study focused specifically on RNA-based prediction, but additional data modalities (*e*.*g*., digital pathology, DNA) might allow for expansion of the diagnostic label and reportable range of the assay. This is particularly relevant in the context of cancer subtypes with pathognomonic alterations such as *BRD4*-*NUT* fusions in NUT midline carcinoma.

Finally, some cancer subtypes can transition from one histological diagnosis to another. For example, prostate and lung adenocarcinomas can acquire neuroendocrine histology in response to treatment with ADT ^47^ and EGFR inhibitors ^48^, respectively. It can be challenging to assign a precise diagnosis in situations where a tumor has a mixed or transitioning histology. Tempus TO identifies the prominent histological subtype in a tumor specimen, enabling better clinical management.

## Conclusion

In summary, the present study has demonstrated the high accuracy and granularity of a cancer subtype predictor (Tempus TO) for use in classifying cancers of unknown or uncertain primary origin. The subtype predictor was built using one of the largest known collections of paired RNA-seq and subtype labels and distinguishes between 68 subtypes with an overall accuracy of 91%. We anticipate that Tempus TO will be a useful tool for providing physicians with a precise histological and site-specific diagnosis, an essential component for clinical decision-making in the increasingly detailed landscape of precision medicine.

## Supporting information

Combined Supplemental Information

All Tables (Main and Supplemental)

## Data Availability

Raw data for this study were generated at Tempus Labs. Derived data supporting the findings of this study are available within the paper and its Supplementary Figures/Tables or available from the authors upon request.

## Notes

### Competing Interest Statement

All authors are employees, former employees, and/or hold restricted stock units of Tempus Labs, Inc.

### Funding Statement

All authors are current or former employees of Tempus Labs, who provided funding for this work.

