## Supplementary material for "Validation of a transcriptome-based assay for classifying cancers of unknown primary origin": Combined Supplemental Information

### Supplementary Methods

#### RNA Normalization

Raw transcript level counts were generated with Kallisto (v0.44.0). Transcripts were quantile-normalized with 100 quantiles to account for GC bias and transcript length.

```
factor = gc_content
global_median = median(expression_vector)
quantile_bins = expression.groupby(factor)
for bin in quantile_bins:
      bin_median = median(bin)
      scaling_factor = bin_median/global_median
      expression = bin/scaling_factor
```

The library size normalization consists of a fit and transform step. During the fit step, a cohort of training data is used to calculate the transcript-wise geometric expression. During the transform step, the learned geometric means are used to calculate a per-sample scaling factor based on the median of the ratio of sample expression to geometric mean of expression.

```
fit:

expression_matrix = [samples x transcripts matrix]
geomean = {}
for transcript in expression_matrix:
      geomean[transcript] = geomean(transcript)

transform:
for sample in expression_matrix:
      scaling_factor = median(sample/geomean)
      sample = sample/scaling_factor
```

#### Batch Correction

The Tempus TO algorithm was trained on a cohort of RNA samples acquired on two different assay versions. Prior to Oct. 2020, samples were run on the version one RNA assay (RS.v1) whereas samples processed after Oct. 2020 were run on version two of the RNA assay (RS.v2). While the differences between assay versions were small in most cases, a simple batch correction method was developed and applied to further reduce differences between assay versions. This method, dubbed “cognizant correction”, leverages ~500 paired samples that were run on both platforms to match the means and variances of normalized expression. Cognizant correction learns per-gene linear transformations to match the RS.v2 data to the RS.v1 data. The slope and intercept coefficients were fitted independently for each gene, using the corresponding normalized expression data, to match their distributions while accounting for pair-membership metadata. Cognizant correction optimizes coefficients by minimizing a loss function based on a weighted version of the Kullback-Leibler divergence that can account for subset-membership metadata, in this case the pairing information.

### Supplementary Figures and Tables

**Figure S1**: Validation of expression with orthogonal qPCR assay. Mean delta CT values for each sample and gene were normalized to the mean of the average CT values of two the housekeeping genes *AAMP* and *CANX* for that sample. Expression data were normalized to control for library size, GC content and transcript length before being log-transformed.

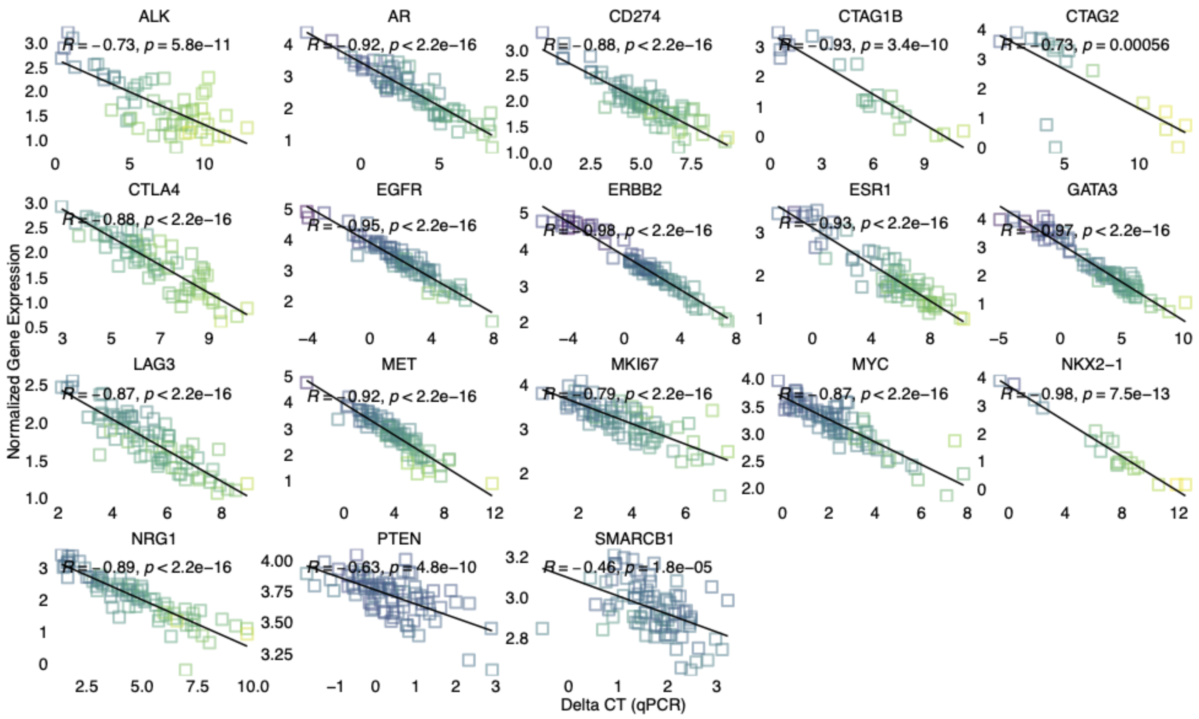

**Figure S2**. Confusion matrices of representative subtype groupings. Row counts refer to true labels, while column counts refer to predicted subtypes. Colors are normalized by the total number of observed counts per subtype, so that high prevalence subtypes (e.g., colorectal adenocarcinoma) can be compared to low prevalence subtypes (e.g., goblet cell adenocarcinoma).

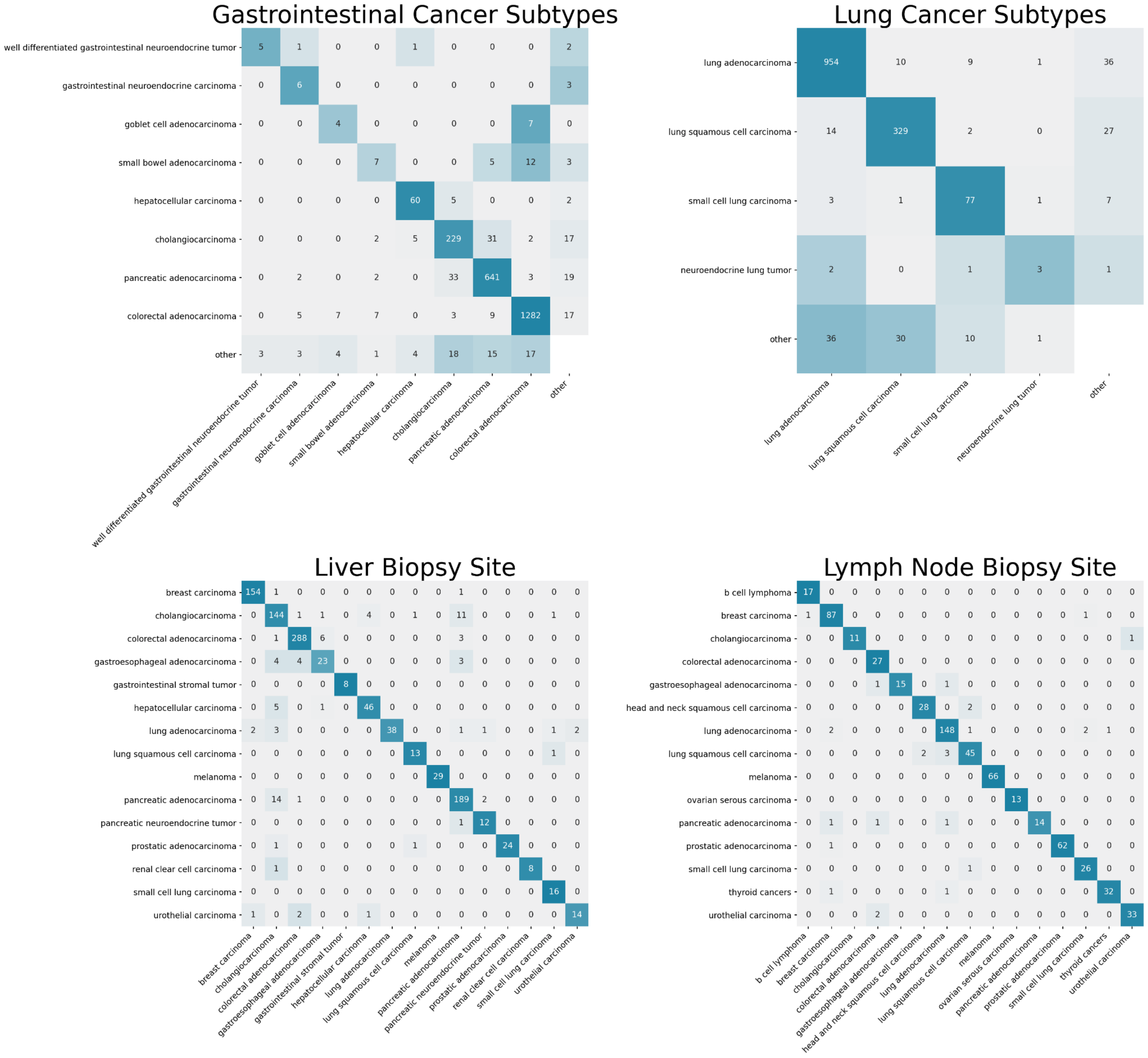

**Figure S3**. Model-predicted (Y-axis) and assigned (X-axis) subtype labels of Tempus samples are evaluated using a confusion matrix.

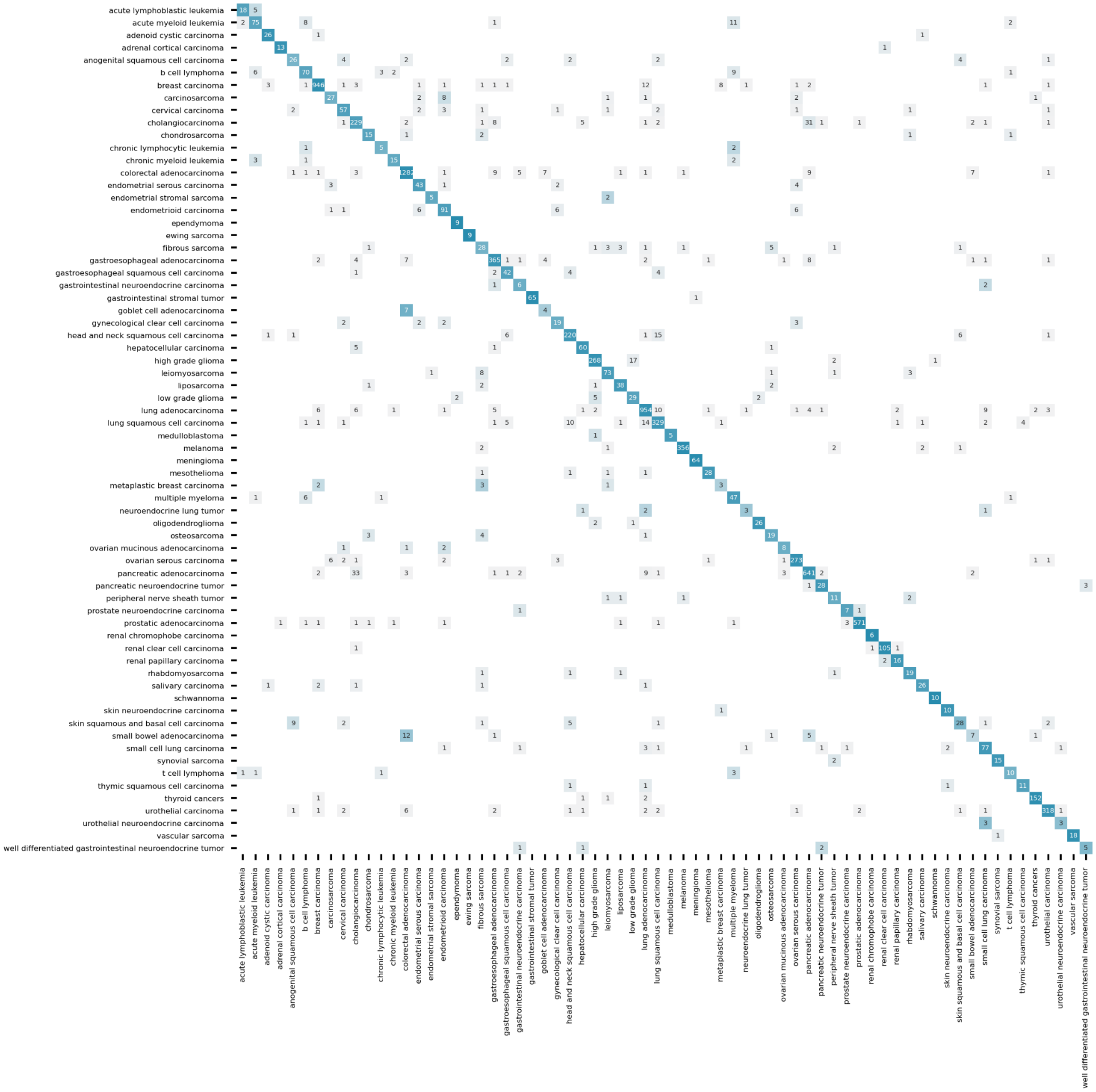

**Figure S4**. Model-predicted (Y-axis) and assigned (X-axis) subtype labels of TCGA samples are evaluated using a confusion matrix.

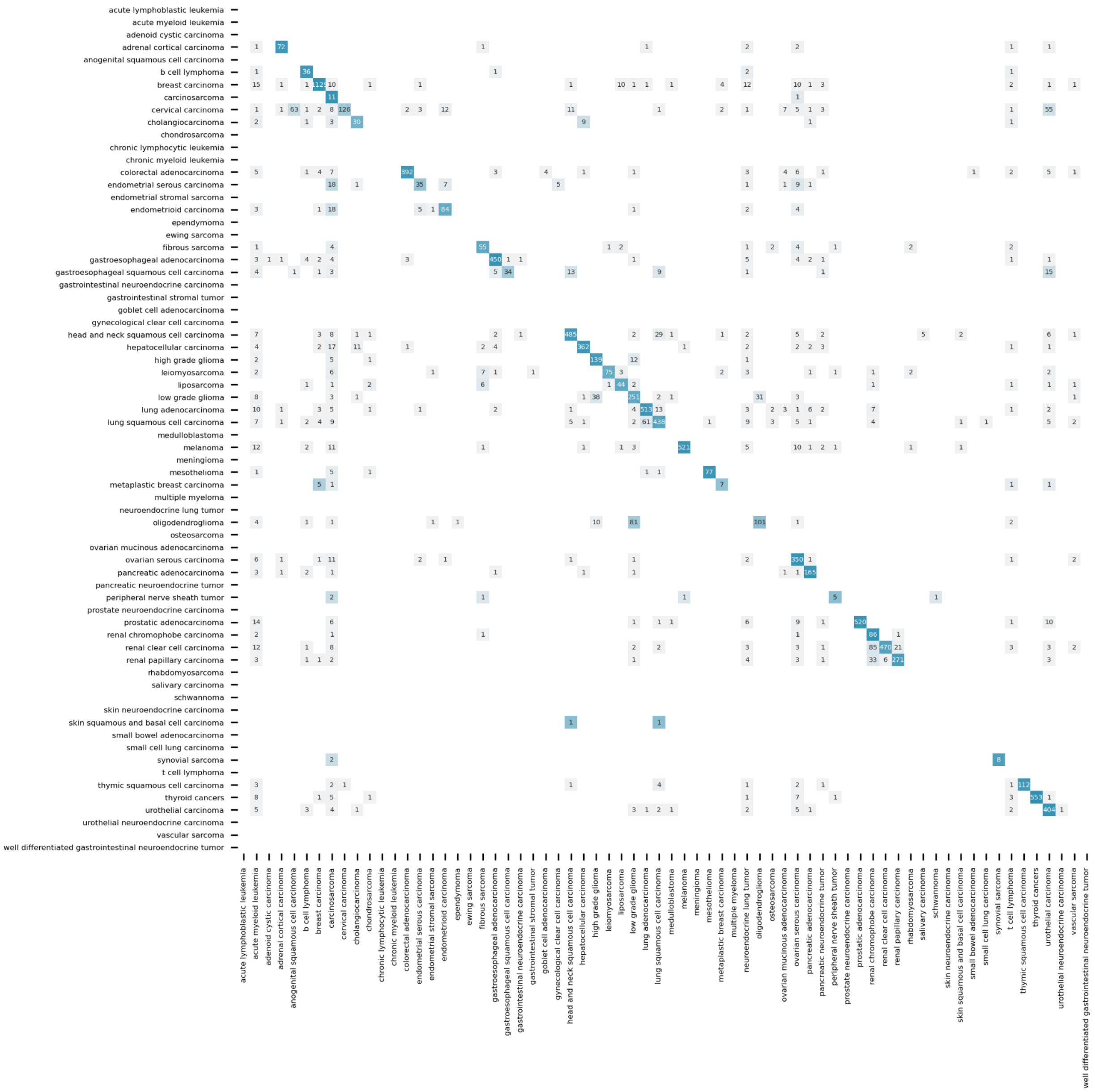

**Table S1**.  Tissue site distribution of training and validation sets.

(See full table in supporting spreadsheet.)

**Table S2**. Subtype distribution of training and validation sets.

(See full table in supporting spreadsheet.)

**Table S3**. Model performance for each cancer subtype and each subcohort (validation, prospective, and retrospective).

(See full table in supporting spreadsheet.)

**Table S4.** A histology-level mapping between TCGA and Tempus TO subtypes is provided.

(See full table in supporting spreadsheet.)

**Table S5**. A crosswalk was constructed to align the subtypes herein to the types in the TCGA dataset.

(See full table in supporting spreadsheet.)

**Table S6**. Model performance on the total validation set and two sub-cohorts (prospective samples that were sequenced after model development and retrospective samples that were randomly held out from model training). Fisher's exact test (p=0.621) was used to assess significance between the cohorts (prospective vs retrospective) and prediction status (correct vs incorrect). Detailed results per subtype are shown in **Table S3**. In the smaller prospective cohort, some subtypes were not observed and were thus excluded from the mean sensitivity calculation.

| | **Validation** | **Prospective** | **Retrospective** |
|---|---|---|---|
| Sample Size | 9210 | 2483 | 6727 |
| Accuracy | 91.1% | 90.8% | 91.2% |
| Top 3 Accuracy | 97.5% | 97.6% | 97.5% |
| Mean Sensitivity | 80.0% | 76.8% | 81.0% |

**Table S7**. Model performance stratified by tumor purity.

| Tumor Purity | N | Sensitivity |
|---|---|---|
| 0.0 < x <= 10.0 | 41 | 80.5% |
| 10.0 < x <= 20.0 | 734 | 89.0% |
| 20.0 < x <= 30.0 | 626 | 90.1% |
| 30.0 < x <= 40.0 | 1229 | 92.9% |
| 40.0 < x <= 50.0 | 1415 | 93.0% |
| 50.0 < x <= 60.0 | 1340 | 92.8% |
| 60.0 < x <= 70.0 | 1332 | 93.0% |
| 70.0 < x <= 80.0 | 1531 | 89.9% |
| 80.0 < x <= 90.0 | 713 | 90.3% |
| 90.0 < x <= 100.0 | 1 | 100.0% |

**Table S8**. Model performance stratified by imputed metastatic status.

| Sample Type | Count | Accuracy |
|---|---|---|
| Ambiguous - Lymph Node Sample | 652 | 91.2% |
| Evidence Against Primary Sample | 359 | 89.5% |
| Possible Metastatic Sample | 1796 | 88.0% |
| Possible Primary Sample | 3041 | 93.5% |
| Possible Recurrent Sample | 335 | 91.0% |
| Unknown | 2204 | 90.6% |

**Table S9**. Model performance on TCGA data is summarized via several performance metrics and descriptive statistics.

| Metric | Value |
|---|---|
| Sample Size | 9976 |
| Accuracy | 84.3% |
| Top 3 Accuracy | 91.4% |
| Mean Sensitivity | 85.2% |

**Table S10**. Per-subtype model performance on TCGA is summarized.

| | observed | tp | sensitivity |
|---|---|---|---|
| acute lymphoblastic leukemia | 0 | 0 | |
| acute myeloid leukemia | 0 | 0 | |
| adenoid cystic carcinoma | 0 | 0 | |
| adrenal cortical carcinoma | 81 | 72 | 88.9% |
| anogenital squamous cell carcinoma | 0 | 0 | |
| b cell lymphoma | 41 | 36 | 87.8% |
| breast carcinoma | 1206 | 1129 | 93.6% |
| carcinosarcoma | 12 | 11 | 91.7% |
| cervical carcinoma | 306 | 126 | 41.2% |
| cholangiocarcinoma | 47 | 30 | 63.8% |
| chondrosarcoma | 0 | 0 | |
| chronic lymphocytic leukemia | 0 | 0 | |
| chronic myeloid leukemia | 0 | 0 | |
| colorectal adenocarcinoma | 441 | 392 | 88.9% |
| endometrial serous carcinoma | 78 | 35 | 44.9% |
| endometrial stromal sarcoma | 0 | 0 | |
| endometrioid carcinoma | 119 | 84 | 70.6% |
| ependymoma | 0 | 0 | |
| ewing sarcoma | 0 | 0 | |
| fibrous sarcoma | 75 | 55 | 73.3% |
| gastroesophageal adenocarcinoma | 485 | 450 | 92.8% |
| gastroesophageal squamous cell carcinoma | 87 | 34 | 39.1% |
| gastrointestinal neuroendocrine carcinoma | 0 | 0 | |
| gastrointestinal stromal tumor | 0 | 0 | |
| goblet cell adenocarcinoma | 0 | 0 | |
| gynecological clear cell carcinoma | 0 | 0 | |
| head and neck squamous cell carcinoma | 564 | 485 | 86.0% |
| hepatocellular carcinoma | 415 | 362 | 87.2% |
| high grade glioma | 160 | 139 | 86.9% |
| leiomyosarcoma | 108 | 75 | 69.4% |
| liposarcoma | 61 | 44 | 72.1% |
| low grade glioma | 340 | 251 | 73.8% |
| lung adenocarcinoma | 581 | 513 | 88.3% |
| lung squamous cell carcinoma | 562 | 438 | 77.9% |
| medulloblastoma | 0 | 0 | |
| melanoma | 573 | 521 | 90.9% |
| meningioma | 0 | 0 | |

| | | | |
|---|---|---|---|
| mesothelioma | 86 | 77 | 89.5% |
| metaplastic breast carcinoma | 15 | 7 | 46.7% |
| multiple myeloma | 0 | 0 | |
| neuroendocrine lung tumor | 0 | 0 | |
| oligodendroglioma | 203 | 101 | 49.8% |
| osteosarcoma | 0 | 0 | |
| ovarian mucinous adenocarcinoma | 0 | 0 | |
| ovarian serous carcinoma | 380 | 350 | 92.1% |
| pancreatic adenocarcinoma | 177 | 165 | 93.2% |
| pancreatic neuroendocrine tumor | 0 | 0 | |
| peripheral nerve sheath tumor | 10 | 5 | 50.0% |
| prostate neuroendocrine carcinoma | 0 | 0 | |
| prostatic adenocarcinoma | 570 | 520 | 91.2% |
| renal chromophobe carcinoma | 92 | 86 | 93.5% |
| renal clear cell carcinoma | 616 | 470 | 76.3% |
| renal papillary carcinoma | 329 | 271 | 82.4% |
| rhabdomyosarcoma | 0 | 0 | |
| salivary carcinoma | 0 | 0 | |
| schwannoma | 0 | 0 | |
| skin neuroendocrine carcinoma | 0 | 0 | |
| skin squamous and basal cell carcinoma | 2 | 0 | 0.0% |
| small bowel adenocarcinoma | 0 | 0 | |
| small cell lung carcinoma | 0 | 0 | |
| synovial sarcoma | 10 | 8 | 80.0% |
| t cell lymphoma | 0 | 0 | |
| thymic squamous cell carcinoma | 128 | 112 | 87.5% |
| thyroid cancers | 581 | 553 | 95.2% |
| urothelial carcinoma | 435 | 404 | 92.9% |
| urothelial neuroendocrine carcinoma | 0 | 0 | |
| vascular sarcoma | 0 | 0 | |
| well differentiated gastrointestinal neuroendocrine tumor | 0 | 0 | |

**Table S11**.  Observed somatic mutation frequency (in the labeled and CUP cohorts) and Fisher p-values for the DNA variant analysis.

(See full table in supporting spreadsheet.)
